# Lithium treatment after electroconvulsive therapy in bipolar disorder: A nationwide target trial emulation

**DOI:** 10.64898/2026.02.11.26346116

**Authors:** Christopher Rohde, Søren Dinesen Østergaard

## Abstract

**Objectives:** Electroconvulsive Therapy (ECT) is an effective treatment for bipolar disorder, particularly in severe acute cases or for illness resistant to pharmacotherapy. However, the risk of relapse following ECT is high, necessitating intervention to reduce this risk. Based on findings from ECT studies in unipolar depression and its well-known mood-stabilizing properties, it is likely that lithium treatment may reduce the risk of relapse of bipolar disorder following ECT.

Therefore, we conducted a target trial emulation using data from Danish nationwide registers to investigate whether lithium protects against relapse following ECT treatment of bipolar disorder.

**Methods:** Patients discharged from their first psychiatric admission with a primary diagnosis of bipolar disorder between January 1, 2006, and June 1, 2024, who received at least six ECT treatments, were included. Follow-up began two weeks after discharge and continued until relapse, death, one year, or January 1, 2025. Patients were considered allocated to lithium treatment if they redeemed a prescription for lithium within the first two weeks after discharge from the index admission (ECT treatment). The outcome was time to relapse, defined by either psychiatric hospital admission or suicide. Cox proportional hazards regression, adjusted for potential confounders, was used to compare the outcome between patients allocated and not allocated to lithium treatment.

**Results:** Among the 574 eligible patients (mean age 41.5 years, 61.3% women), 214 (37.3%) were allocated to lithium treatment and 360 (62.7%) were not allocated to lithium treatment. During follow-up, 56 patients (26.2%) in the lithium group and 135 patients (37.5%) in the non-lithium group experienced a relapse. Lithium treatment was associated with a substantially reduced risk of relapse (adjusted hazard rate ratio, 0.60, 95% CI=0.43-0.84).

**Conclusion:** Lithium treatment after ECT may reduce the risk of relapse in patients with bipolar disorder. These findings should be followed up by a randomized controlled trial.

## 1. INTRODUCTION

Electroconvulsive Therapy (ECT) is widely acknowledged as one of the most effective treatments for patients with bipolar disorder, irrespective of the type of mood episode.^1-4^ However, the duration of the response is often limited, with many patients experiencing relapse within a relatively short period.^5-7^ This high risk of relapse poses a clinical challenge, highlighting the need for optimal mood-stabilizing treatment following ECT. Lithium is particularly interesting in this regard due to its ability to reduce the frequency and severity of episodes of bipolar disorder and decrease the risk of suicide.^8-12^

In a recent systematic review and meta-analysis, Lambrichts et al. found that the available clinical and epidemiological evidence (14 studies in total) suggests that patients with depression who receive lithium following ECT are less likely to experience relapse compared with those not receiving lithium.^13^ Seven of the studies in this review included patients with bipolar disorder.^7,14-19^ However, in the six studies comprising both patients with unipolar depression and bipolar disorder, the latter representing the minority, analyses focusing exclusively on the fraction of patients with bipolar disorder were not conducted. Only the register-based study by Popiolek et al. ^7^ specifically investigated whether lithium was associated with decreased risk of relapse following ECT in bipolar depression. This study found that treatment with lithium treatment after ECT for bipolar depression was linked to reduced risk of relapse, which was, albeit not statistically significant. However, this study did not exclude patients who used lithium prior to receiving ECT,^7^ potentially introducing a prevalent-user bias.^20^ Indeed, lithium treatment prior to ECT was associated with an increased risk of relapse following ECT.^7^

Here, we used nationwide Danish register data to conduct a target trial emulation investigating the effect of lithium on the risk of relapse following ECT treatment of bipolar disorder. A target trial emulation refers to the emulation of a hypothetical randomized clinical trial that may not be feasible to conduct in practice, for instance by using data from population-based registers.^21-24^ We defined the target trial (i.e., the trial we sought to emulate), adhering to the principles established by Hernan et al.^21^ that specifically counter prevalent-user bias by aligning the start of exposure (lithium treatment) with the start of follow-up.

## 2. METHODS

### 2.1. Protocol and approvals

The target trial protocol was preregistered on the Open Science Framework: https://osf.io/9hu5g. We followed the protocol with two exceptions, namely that we i) used data up until January 1, 2025 (instead of until March 1, 2019), as we received a data update following the preregistration, and ii) we only received data on suicide (part of the operationalization of relapse) up until December 31, 2022. Consequently, suicides beyond that date were not included as outcomes.

The Danish Health Data Authority and Statistics Denmark approved the use of register data for the purpose of this study. Register-based studies are exempt from ethical review board approval according to Danish law (waiver no. 1-10-72-116-25). The study was registered on the internal list of research projects having Aarhus University as data steward.

### 2.2. Target Trial Specification

First, we specified the hypothetical trial we sought to emulate (the target trial) as shown in Table 1 (with the updated end of data: January 1, 2025). This target trial would include patients discharged from their first psychiatric admission (index admission) with bipolar disorder as the main diagnosis (International Classification of Diseases, 10th Revision [ICD-10] codes: F30, F31) and who received a series of ECT (a minimum of 6 ECT treatments) during this admission in the period from January 1, 2006 to June 1, 2024. The trial would assess eligibility at the discharge date, with exclusion criteria as follows: i) Age <18 years at discharge, ii) a prior diagnosis of schizophrenia (ICD-10 code: F20) or schizoaffective disorder (ICD-10 code: F25), and iii) prior use of lithium – to avoid prevalent-user bias. In the two weeks following discharge, the target trial would involve random assignment of the included patients to one of two open-label treatment arms: i) treatment with lithium or ii) no treatment with lithium. Follow-up would begin two weeks after discharge and continue until relapse (psychiatric admission or suicide), death, one year, or January 1, 2025 (end of data), whichever occurred first. Patients readmitted within two weeks of discharge would be excluded, as a readmission this close to the discharge date would be more likely to be due to residual symptoms from the initial admission than a relapse. The primary outcome of the target trial would be relapse. As in our recent target-trial emulation,^24^ the outcome comparisons between the two treatment arms would be analyzed through an intention-to-treat (ITT) approach using Cox proportional hazards regression.

**Table 1.** Target trial emulation protocol.

| Protocol Component | Target Trial specification | Target Trial Emulation |
| --- | --- | --- |
| Eligibility criteria | 1. Discharged from the first psychiatric admission (index) with bipolar disorder as the main diagnosis (ICD-10: F30, F31) between January 1, 2006 and June 1, 2024*.<br>2. Aged $\geq 18$ years when discharged from the index admission.<br>3. Have received a series of electroconvulsive therapy (ECT) treatments (a minimum of 6 ECT treatments) during the index admission.<br>4. No prior diagnosis (any type) of schizophrenia (ICD-10: F20) or schizoaffective disorder (ICD-10: F25).<br>5. No previous use of lithium. | Same as for the target trial. |
| Treatment strategies | 1. Treatment with lithium<br>2. No treatment with lithium | Same as for the target trial. Patients will be identified as treated with lithium if redeeming a prescription for lithium within the first two weeks after discharge from the index admission. |
| Assignment | Patients are randomly assigned to the treatment strategy at baseline. Individuals are aware of their assigned treatment. | We assume randomization conditional on the following baseline covariates (all assessed in the two years leading up to the index admission date): age, sex, calendar year, educational level, marital status, occupational status, somatic comorbidity (the Charlson Comorbidity index), use of a sedative/hypnotic drug, use of a mood-stabilizing drug (antipsychotic, valproate or lamotrigine), use of an antidepressant, the severity of bipolar disorder during the index admission (hypomania/non-psychotic mania [F30.0, F30.1, F30.8, F30.9 and F31.0-F31.1], non-psychotic bipolar depression [F31.3-F31.4], psychotic mania, psychotic depression or mixed episode [F30.2, F31.2, F31.5 or F31.6], and other bipolar episodes [F31.7-F31.9]), psychotic comorbidity (diagnosis of psychotic disorder excl. schizophrenia and schizoaffective disorder [F2X excl. F20 and F25]), non-psychotic comorbidity (diagnosis of substance use disorder [F10-19], anxiety/OCD [F40, F41, F42], personality disorder [F60, F61], or ADHD [F90]), and prior psychiatric admission (yes/no). |
| Follow-up | For eligible patients, follow-up starts two weeks after discharge and continues until:<br>• Any psychiatric admission<br>• Death<br>• End of follow-up (January 1, 2025*)<br>• One year after discharge<br>- whichever occurs first. Patients that are readmitted within 2 weeks after discharge following the index admission are excluded. | Same as for the target trial. |
| Outcomes | Any psychiatric admission or suicide** (merged) | Same as for the target trial |
| Causal contrasts | Intention-to-treat effect. | Observational analogue of the intention-to-treat effect |
| Data analysis | Intention-to-treat analysis. | Same as for the target trial, via Cox proportional hazards regression adjusted for the covariates listed under “Assignment”. |

\* In the preregistered protocol published on the Open Science Framework (https://osf.io/9hu5g), this date is March 1, 2019.
\*\* As we only had data on suicide until December 31, 2022, suicides beyond this date could not be included as outcomes.

**Table 2.** Baseline clinical and demographic characteristics of the individuals included in the target trial emulation.

|  | Lithium treatment<br>(N = 214) | No lithium treatment<br>(N = 360) |
| --- | --- | --- |
| Females, n (%) | 123 (57.5%) | 229 (63.6%) |
| Age at time of inclusion, mean (SD) | 46.2 (16.9) | 54.6 (17.8) |
| Calendar year, n (%) |  |  |
| <2012 | 65 (30.4%) | 149 (41.4%) |
| 2012-2015 | 38 (17.8%) | 78 (21.7%) |
| 2016-2020 | 62 (29.0%) | 92 (25.6%) |
| 2021-2025 | 49 (22.9%) | 41 (11.4%) |
| Bipolar disorder severity, n (%) |  |  |
| Hypomania/non-psychotic mania | 25 (11.7%) | 26 (7.2%) |
| Non-psychotic depression | 61 (28.5%) | 123 (34.2%) |
| Psychotic mania, psychotic depression, or mixed episode | 99 (46.3%) | 149 (41.4%) |
| Other bipolar episode | 29 (13.6%) | 62 (17.2%) |
| Psychiatric comorbidity, n (%) |  |  |
| Psychotic comorbidity | 27 (12.6%) | 40 (11.1%) |
| Non-psychotic comorbidity | 49 (22.9%) | 88 (24.4%) |
| Prior psychiatric inpatient admission | 95 (44.4%) | 158 (43.9%) |
| Prior psychopharmacological treatment, n (%) |  |  |
| Mood-stabilizing agent | 116 (54.2%) | 223 (61.9%) |
| Sedative/hypnotic | 114 (53.3%) | 222 (61.7%) |
| Antidepressant | 157 (73.4%) | 281 (78.1%) |
| Charlson comorbidity index, n (%) |  |  |
| 0 | 194 (90.7%) | 297 (82.5%) |
| >=1 | 20 (9.3%) | 63 (17.5%) |
| Civil status, n (%) |  |  |
| Living with a partner | 85 (39.7%) | 144 (40.0%) |
| Not living with a partner | 129 (60.3%) | 216 (60.0%) |
| Education level, n (%) |  |  |
| No education or primary and lower secondary school | 59 (27.6%) | 121 (33.6%) |
| High school/vocational school | 74 (34.6%) | 138 (38.3%) |
| Short- or medium-term higher education | 57 (26.6%) | 77 (21.4%) |
| Long-term higher education | 24 (11.2%) | 24 (6.7%) |
| Current employment status, n (%) |  |  |
| Employed | 85 (39.7%) | 105 (29.2%) |
| Unemployed | 77 (36.0%) | 90 (25.0%) |
| Outside the labour force | 52 (24.3%) | 165 (45.8%) |

### 2.3. Target Trial Emulation

The target trial was emulated (see Table 1) using observational data from nationwide Danish registers.^25^ Specifically, we obtained information on birthplace, date of birth, and vital status from the Danish Civil Registration System (DCRS).^26^ The DCRS includes a unique personal identifier allowing linkage across multiple registers for all residents of Denmark. Data on contacts with Danish psychiatric hospitals, including admissions, outpatient contacts, emergency visits and ECT were obtained from the Danish National Patient Register (DNPatR).^27^ Notably, on March 1, 2019, the DNPatR changed from the second to the third version.^28^ In the third version, contacts are no longer specified as inpatient or outpatient contacts. From March 1, 2019, we, therefore, defined contacts with psychiatric services lasting more than 12 hours as admissions.^29^ Data on redeemed prescriptions were obtained from the Danish National Prescription Register (DNPreR).^30^ Data on death and suicide (ICD-10 code for cause of death: X60-X84, Y870) were obtained from the Danish Cause of Death Register.^31^ Data on the highest obtained level of education and employment status were obtained from the Population Education Register and Employment Classification Module ^32^, respectively. All data mentioned above are time-stamped.

#### Study population: Patients with bipolar disorder receiving ECT

Using data from the DPCRR and DNPatR, we identified patients discharged from their first psychiatric admission with bipolar disorder (ICD-10: F30, F31) as the main diagnosis and who received a series of ECT (a minimum of 6 ECT treatments) during this admission in the period from January 1, 2006, to June 1, 2024.

We excluded patients younger than 18 years at discharge, those with a prior diagnosis of schizophrenia or schizoaffective disorder (from January 1, 1994 – the implementation date of ICD-10 in Denmark – and onwards), and those with a history of lithium use (from January 1, 1995 – the beginning of the DNPreR – and onwards).

#### Intervention (lithium treatment)

In line with our recent target trial emulation,^24^ patients were identified as allocated to lithium treatment if they redeemed a prescription for lithium (Anatomical Therapeutic Chemical (ATC): N06A) within the first two weeks after discharge from the index admission. Other patients were considered not allocated to lithium treatment.

#### Outcomes

The primary outcome was relapse, defined as psychiatric admission or suicide (as in Popiolek et al. ^7^) within the follow-up period. Since we only had data on suicide up until December 31, 2022, suicides beyond this date could not be included as outcomes.

#### Follow-up

We followed all included patients from two weeks post-discharge until the first occurrence of relapse, death, one year after initiation of follow-up, or end of data (January 1, 2025). Patients admitted or who died within the initial two weeks post-discharge were excluded from the analysis.

### 2.4. Statistical Analysis

The outcome rate for the two treatment groups was compared via Cox proportional hazards regression, focusing on the intention-to-treat effect. We adjusted for baseline covariates to emulate the conditions of random treatment allocation, assuming randomization conditional on the following baseline covariates: age, sex, calendar year, educational level (primary school, high school or further education), marital status (living with partner, not living with partner), occupational status (employed, non-employed, or retired), medical comorbidity (number of Charlson Comorbidity index diseases, 0 or ≥1), use of sedatives/hypnotics, use of mood-stabilizing drugs other than lithium (valproate, lamotrigine or an antipsychotic), use of antidepressants, type of mood episode during the index admission (categorized as non-psychotic mania, non-psychotic bipolar depression, psychotic mania, psychotic depression or mixed episode, and other bipolar episodes), psychotic comorbidity (other primary psychotic disorder than schizophrenia and schizoaffective disorder), non-psychotic comorbidity (substance use disorder, anxiety/OCD, personality disorder, or ADHD), and prior psychiatric admission (yes/no). For the operationalization of these disease/disorder- and drug covariates using ICD-10 and Anatomical Therapeutic Chemical classification codes, respectively, see Supplementary Table 1. The covariates were created based on information from the two years preceding the psychiatric admission.

We visualized the cumulative incidence of relapse for the two treatment groups using Aalen-Johansen curves. We compared the risk of relapse for the two treatment groups using a Cox proportional hazard model adjusted for the covariates mentioned above, yielding adjusted hazard rate ratios (aHRR). The proportional hazards assumption was examined by visual inspection of the log-log survival functions. All statistical analyses were conducted using STATA (version 19).

#### Causal contrast analyses

As patients in the lithium group could stop using lithium during follow-up and patients in the non-lithium groups could start using lithium during follow-up, we conducted analyses to estimate the causal contrast of the study. First, we investigated the level of treatment adherence in the lithium group by calculating the medication possession ratio (MPR) of lithium ^33^ as follows: MPR = ((lithium dose × number of lithium pills per package/Defined Daily Dosage (DDD) / (number of days in observation period)) × 100. Second, we calculated the proportion of patients in the non-lithium group who redeemed a prescription of lithium during follow-up. Third, for those in the non-lithium group redeeming a lithium prescription, we calculated the median time from the start of follow-up to the first redemption of a lithium prescription. In all three analyses, we only included patients with an index date before June 1, 2023, to ensure that all patients had at least one year of follow-up.

#### Post-hoc analyses

First, as 19.7% of patients in the non-lithium group did not redeem a prescription for a mood-stabilizer or an antidepressant in the two weeks following discharge, we repeated the analyses excluding these patients. Second, because the potential protective effect of lithium on relapse in bipolar disorder may depend on mood episode polarity ^34^, we repeated the analyses, restricting to patients with index admission for hypomania/mania (ICD-10: F30.0, F30.1, F30.2, F30.8, F30.9, F31.0, F31.1, F31.2) or for depression (F31.3, F31.4, F31.5), respectively. Third, we repeated the analyses, restricting to patients who had not redeemed a prescription for a mood-stabilizing agent (same as above) in the two years leading up to the index admission.

#### Prior analysis

In 2024, the analysis was run using data up until March 1, 2019. The results of this analysis have not been published but were shown at the 26^th^ annual conference of the International Society of Bipolar Disorders.^35^ We were awaiting the data update at the time.

## 3. RESULTS

### Study population

Figure 1 shows the recruitment and treatment allocation for the target trial emulation. We identified 574 patients fulfilling the eligibility criteria. A total of 214 (37.2%) of these received lithium treatment.

**Figure 1.**
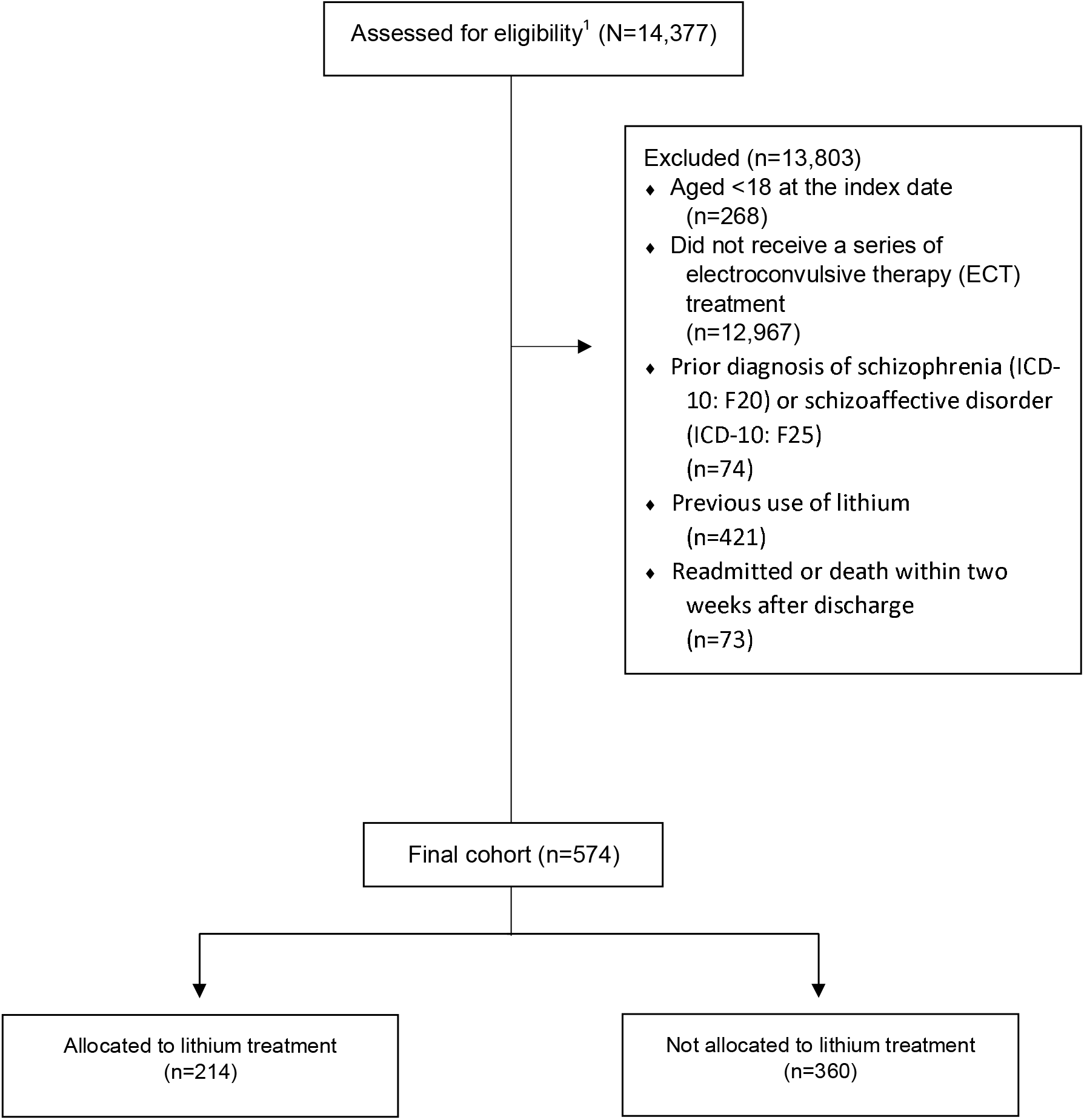
Flowchart showing the selection of the study population ^1^ Discharged from the first psychiatric admission (index) with bipolar disorder as the main diagnosis (ICD-10: F30, F31) between January 1, 2006, and June 1, 2024.

Table 1 lists the characteristics of the patients included in the target trial emulation. Those allocated to lithium treatment tended to be younger, have a longer education, be employed, and have less medical comorbidity than patients not allocated to lithium treatment.

Among patients allocated to lithium treatment, 21 (9.8%) received (i.e., redeemed a prescription) lithium and an antidepressant, 122 (57.0%) received lithium and another mood-stabilizing agent, 49 (22.9%) received lithium, an antidepressant, and a mood-stabilizing agent, and 22 (10.3%) received lithium monotherapy. Among patients not allocated to lithium treatment, 29 (8.1%) received antidepressant monotherapy, 134 (37.2%) received a mood-stabilizing agent, 126 (35.0%) received an antidepressant and a mood-stabilizing agent, and 71 (19.7%) did not redeem any prescription of the medications mentioned above in the two weeks following the discharge.

### Comparison of relapse rates

Table 3 lists the incidence rate of relapse during follow-up and the aHRR comparing those receiving lithium to those not receiving lithium, while Figure 2 shows the cumulative incidence of relapse in the two groups. All relapses were psychiatric admissions, as there were no suicides during follow-up. During follow-up, 56 patients (26.2%) in the lithium group and 135 patients (37.5%) in the non-lithium group experienced a relapse. Lithium treatment was associated with a lower risk of relapse (aHRR=0.60, 95%CI=0.43-0.84, p<0.01). Similar results were found when males and females were analyzed separately (Table 3). The proportional hazards assumption was met (see Supplementary Figure 1).

**Figure 2.**
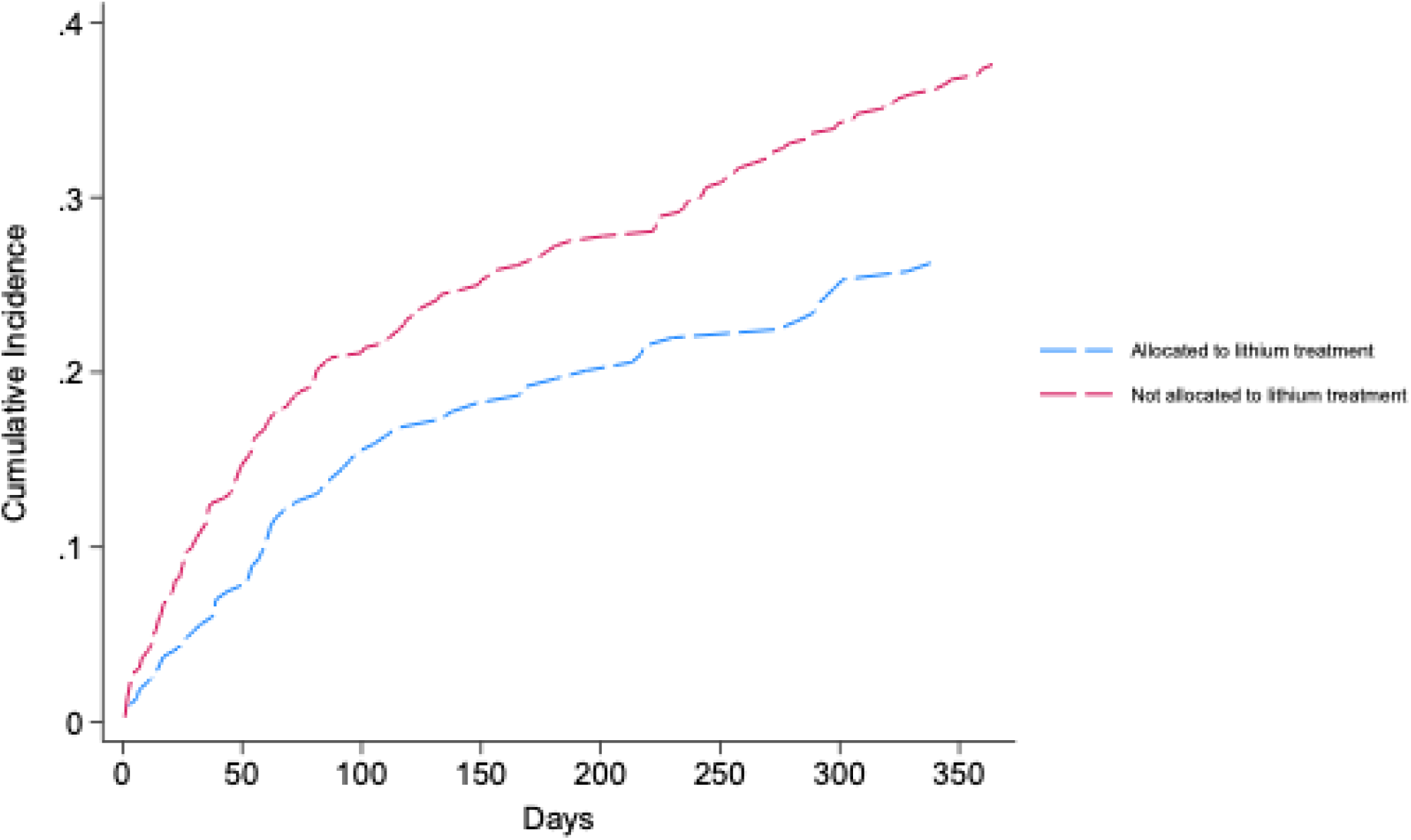
Cumulative incidence proportion (relapse)

**Table 3.** Risk of psychiatric admission or suicide in patients with bipolar disorder treated with ECT allocated to lithium treatment vs. no lithium treatment.

|  |  |  | Partly adjusted model<sup>b</sup> |  | Fully adjusted model<sup>c</sup> |  |
| --- | --- | --- | --- | --- | --- | --- |
|  | Number of outcomes | Incidence rate (95%CI)<sup>a</sup> | HRR | 95%CI | HRR | 95%CI |
| **All patients (n = 574)** |  |  |  |  |  |  |
| Lithium (n = 214) | 56 | 321.6 (247.5-417.9) | 0.60 | 0.44-0.83<sup>d</sup> | 0.60 | 0.43-0.84<sup>d</sup> |
| No Lithium (n = 360) | 135 | 511.2 (431.9-605.2) | 1.00 (ref.) | - | 1.00 (ref.) | - |
| **Women (n = 352)** |  |  |  |  |  |  |
| Lithium (n = 123) | 34 | 340.8 (243.5-477.0) | 0.64 | 0.42-0.98 | 0.60 | 0.39-0.93 |
| No Lithium (n = 229) | 81 | 474.2 (381.4-589.5) | 1.00 (ref.) | - | 1.00 (ref.) | - |
| **Men (n = 222)** |  |  |  |  |  |  |
| Lithium (n = 91) | 22 | 295.8 (194.8-449.3) | 0.55 | 0.33-0.91 | 0.55 | 0.32-0.95 |
| No Lithium (n = 131) | 54 | 579.2 (443.6-756.3) | 1.00 (ref.) | - | 1.00 (ref.) | - |

<sup>a</sup> Incidence rate per 1000 person-years
<sup>b</sup> Adjusted for age, sex and calendar year.
<sup>c</sup> Adjusted for age, sex, calendar year, educational level, marital status, occupational status, somatic comorbidity (the Charlson Comorbidity index), use of a sedative/hypnotic drug, use of a mood-stabilizing drug, use of an antidepressant, the severity of bipolar disorder during the index admission, psychotic comorbidity, non-psychotic comorbidity, and prior psychiatric admission.
<sup>d</sup> The standard error was 0.100 for the partially adjusted model and 0.102 for the fully adjusted model.
CI = Confidence interval. HRR = hazard rate ratio.

### Causal contrast analyses

The lithium group had a median (25^th^-75^th^ percentile) MPR of 86.8% (51.1-116.4), and 87.9% redeemed another lithium prescription during follow-up. A total of 10.6% of the patients in the non-lithium group redeemed a prescription for lithium during follow-up. For the 10.6% of patients in the non-lithium group redeeming a lithium prescription during follow-up, the median time (25^th^- 75^th^ percentile) from start of follow-up to the first redemption of a lithium prescription was 32 days (11-105).

### Post-hoc analyses

The association between lithium treatment and lower risk of relapse remained consistent across all post-hoc analyses (see Supplementary Tables 2-5).

### Prior analysis

In the analysis based on data up until March 1, 2019 (based on 421 patients with 134 receiving lithium and 287 not receiving lithium), lithium was also associated with a reduced risk of relapse, which was, however, not statistically significant (aHRR=0.74, 95%CI=0.50-1.08).

## 4. DISCUSSION

In this target trial emulation, including 574 patients with bipolar disorder receiving ECT, we found that initiation of lithium treatment following ECT was associated with a substantially reduced risk of relapse. Overall, 26% of patients allocated to lithium treatment experienced a relapse compared with 38% in the non-lithium group. Comparison of hazard rates for relapse, with adjustment for likely confounders, supported that this difference was due to lithium. These findings suggest that mood-stabilizing treatment with lithium should be considered when ECT has been used in bipolar disorder.

While there is a relatively large evidence base from clinical and epidemiological studies supporting the relapse-protecting potential of lithium following ECT for depression in general,^13^ we are only aware of the study by Popiolek et al.,^7^ which has focused specifically on bipolar disorder.

While the results of this study were suggestive of a protective effect of lithium, they did not reach statistical significance and were potentially biased by inclusion of prevalent lithium users.^7^ As the present target trial emulation essentially rules out prevalent user bias, we believe that its results are currently the best available evidence on the relapse-protecting effect of lithium treatment after ECT in bipolar disorder. However, a target trial emulation is not a randomized controlled trial (RCT) and the promising results from the present study should ideally be followed up by an RCT.

There are limitations to this study. First and foremost, residual confounding cannot be fully excluded. We note some differences in baseline covariates between the groups. Patients not receiving lithium treatment had more medical comorbidity and lower socioeconomic status (including lower education levels and a higher proportion being outside the labor force). There was, however, minimal change in the association estimates from the partly to fully adjusted model taking these differences into account, suggesting that residual confounding from related, but unmeasured, variance may not be a pronounced problem. However, residual confounding *by indication* is likely the main threat to causal inference in target trial emulation. In the context of the present study, such confounding could draw the results in opposite directions; One one hand, as lithium treatment requires close monitoring with a blood-testing regimen, clinicians may be more likely to suggest this treatment option to patients with good insight, health literacy and overall functioning (less severe illness) that can comply with the test regimen, and who may also have relatively low risk of relapse. On the other hand, as lithium is considered by some to be the most potent mood-stabilizer, clinicians could also tend to suggest this treatment to patients with more severe illness that have a higher risk of relapse. The findings from Popiolek et al. are indirectly suggestive of the latter, as patients having received lithium prior to ECT were more likely to experience relapse after ECT.^7^ While we did our utmost to adjust for such confounding by indication via inclusion of available proxies for severity (type of mood episode during the index admission, psychopharmacological treatment other than lithium, psychiatric comorbidity, prior psychiatric admission) and functional impairment (socioeconomic status) in the adjustment of our analyses, the registers do not hold more fine-grained data on patient characteristics that may influence treatment decisions, for instance psychometric ratings during the index admission at which ECT was given. However, by deliberately only including patients discharged from their first psychiatric admission with bipolar disorder and who received ECT, we end up with a relatively homogeneous patient group with severe illness at the time of their first admission for bipolar disorder, likely reducing the potential for confounding by indication. Second, this design choice prioritizing homogeneity reduces the generalizability of the findings. Accordingly, future studies could include patients more broadly, e.g., those receiving ECT after several admissions or who start ECT as outpatients. Third, as lithium treatment requires regular clinical monitoring, clinicians may detect emerging symptoms earlier in the lithium group. This could lead to more timely outpatient intervention and thereby prevent progression to a full relapse requiring psychiatric hospitalization or leading to suicide. This potential detection bias might partially contribute to the lower relapse rate observed in the lithium group. On the other hand, patients with bipolar disorder treated with ECT as inpatients are typically followed closely after discharge, irrespective of whether they receive lithium or not, which counters this bias.

In conclusion, the results of this target trial emulation suggest that lithium treatment following ECT reduces the risk of relapse in patients with bipolar disorder. Ideally, these findings should be followed up by an RCT.

## Data Availability Statement

The data cannot be shared due to restrictions enforced by Danish law for protecting patient privacy.

## Author contributions

The study was designed by CR and SDØ. SDØ procured the data. The statistical analyses were carried out by CR. CR and SDØ contributed to the interpretation of the results. CR and SDØ wrote the manuscript in collaboration. Both authors approved the final version of the manuscript prior to submission.

