## Supplementary Material for "Lithium treatment after electroconvulsive therapy in bipolar disorder: A nationwide target trial emulation"

Christopher Rohde, MD PhD, Søren Dinesen Østergaard, MD PhD

**eTable 1.** Definition of variables based on ATC and ICD-10 codes.

| Lithium | ATC: N05AN01 |
| --- | --- |
| Antidepressants | ATC: N06A |
| Antipsychotics | ATC: N05A (excluding N05AN01) |
| Valproate | ATC: N03AG01 |
| Lamotrigine | ATC: N03AX09 |
| Sedatives/hypnotics | ATC: N05BA, N05C |
| Bipolar disorder severity |  |
| Hypomania/non-psychotic mania | ICD-10: F30.0, F30.1, F30.8, F30.9, F31.0, F31.1 |
| Non-psychotic depression | ICD-10: F31.3, F31.4 |
| Psychotic mania, psychotic depression, or mixed episode | ICD-10: F30.2, F31.2, F31.5, F31.6 |
| Other bipolar episode | ICD-10: F31.7, F31.8, F31.9 |
| Psychotic comorbidity | ICD-10: F20-29 (excluding F20, F25) |
| Non-psychotic comorbidity | ICD-10: F10-19, F40, F41, F42, F60, F61, F90 |
| Charlson’ Comorbidity index:  Myocardial infarction  Congestive Heart failure  Peripheral vascular disease  Cerebrovascular disease  Dementia  Chronic pulmonary disease  Connective tissue disease  Ulcer disease  Mild liver disease  Hemiplegia  Moderate to severe renal disease  Any tumor  Leukemia  Lymphoma  Moderate to severe liver disease  Metastatic solid tumor  AIDS | ICD-10: I21-I23  ICD-10: I50, I11.0, I13.0, I13.2  ICD-10: I70-I74, I77  ICD-10: I60-I69, G45, G46  ICD-10: F00-F03, F05.1, G30  ICD-10: J40-J47, J60-J67, J68.4, J70.1, J70.3, J84.1, J92.0, J96.1, J98.2, J98.3  ICD-10: M05, M06, M08, M09, M30-M36, D86  ICD-10: K22.1, K25-K28  ICD-10: B18; K70.0-K70.3; K70.9; K71; K73; K74; K76.0  ICD-10: G81, G82  ICD-10: I12, I13, N00-N05, N07, N11, N14, N17-N19, Q61  ICD-10: C00-C75  ICD-10: C91-C95  ICD-10: C81-C85, C88, C90, C96  ICD-10: B15.0, B16.0, B16.2, B19.0, K70.4, K72, K76.6, I85  ICD-10: C76-C80  ICD-10: B21-B24 |

**eFigure 1.** Log-log plot showing proportional hazards

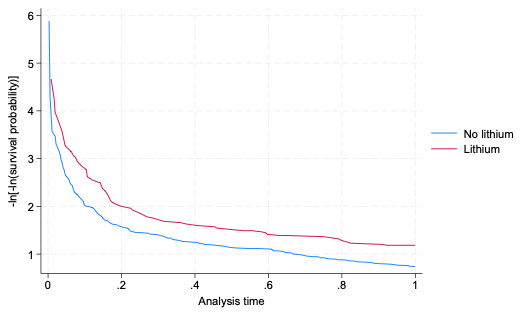

**eTable 2.** Risk of relapse in patients with bipolar disorder treated with ECT allocated to lithium treatment vs. no lithium treatment, excluding the 19.7% that did not redeem a prescription for a mood-stabilizing or antidepressant agent in the non-lithium group.

|  |  |  | **Partly adjusted model^b^** | | **Fully adjusted model^c^** | |
| --- | --- | --- | --- | --- | --- | --- |
|  | **Number of otucomes** | **IR**  **(95%CI)^a^** | **HRR** | **95%CI** | **HRR** | **95%CI** |
| **All patients (n = 503)** | | | | | | |
| Lithium (n = 214) | 56 | 321.6  (247.5-417.9) | 0.53 | 0.38-0.75 | 0.55 | 0.39-0.78 |
| No Lithium  (n = 289) | 116 | 555.5  (463.1-666.4) | 1.00 (ref.) | - | 1.00 (ref.) | - |

**eTable 3.** Risk of relapse in patients with bipolar disorder treated with ECT allocated to lithium treatment vs. no lithium treatment, only including patients with a diagnosis of mania in the index admission.

|  |  |  | **Partly adjusted model^b^** | | **Fully adjusted model^c^** | |
| --- | --- | --- | --- | --- | --- | --- |
|  | **Number of otucomes** | **IR**  **(95%CI)^a^** | **HRR** | **95%CI** | **HRR** | **95%CI** |
| **All patients (n = 117)** | | | | | | |
| Lithium (n = 56) | 12 | 259.5  (147.4-457.0) | 0.68 | 0.32-1.47 | 0.61 | 0.22-1.67 |
| No Lithium  (n = 61) | 18 | 369.0  (232.5-585.7) | 1.00 (ref.) | - | 1.00 (ref.) | - |

**eTable 4.** Risk of relapse in patients with bipolar disorder treated with ECT allocated to lithium treatment vs. no lithium treatment, only including patients with a diagnosis of depression in the index admission.

|  |  |  | **Partly adjusted model^b^** | | **Fully adjusted model^c^** | |
| --- | --- | --- | --- | --- | --- | --- |
|  | **Number of otucomes** | **IR**  **(95%CI)^a^** | **HRR** | **95%CI** | **HRR** | **95%CI** |
| **All patients (n = 291)** | | | | | | |
| Lithium (n = 95) | 26 | 338.7  (230.6-497.5) | 0.62 | 0.39-0.98 | 0.62 | 0.39-0.99 |
| No Lithium  (n = 196) | 73 | 505.5  (401.9-635.9) | 1.00 (ref.) | - | 1.00 (ref.) | - |

**eTable 5.** Risk of relapse in patients with bipolar disorder treated with ECT allocated to lithium treatment vs. no lithium treatment, restricting to patients who had not received a mood-stabilizing agent before the index admission.

|  |  |  | **Partly adjusted model^b^** | | **Fully adjusted model^c^** | |
| --- | --- | --- | --- | --- | --- | --- |
|  | **Number of otucomes** | **IR**  **(95%CI)^a^** | **HRR** | **95%CI** | **HRR** | **95%CI** |
| **All patients (n = 235)** | | | | | | |
| Lithium (n = 98) | 28 | 356.9  (246.5-517.0) | 0.59 | 0.36-0.96 | 0.54 | 0.32-0.90 |
| No Lithium  (n = 137) | 54 | 528.5  (404.7-690.0) | 1.00 (ref.) | - | 1.00 (ref.) | - |
